# SPLUNC1: A Novel Marker of Cystic Fibrosis Exacerbations

**DOI:** 10.1101/2020.05.15.20100669

**Authors:** Sara Khanal, Megan Webster, Naiqian Niu, Myra Nunez, Geoffrey Chupp, Martin D. Slade, Lauren Cohn, Maor Sauler, Jose L. Gomez, Robert Tarran, Lokesh Sharma, Charles S. Dela Cruz, Marie Egan, Theresa Laguna, Clemente J. Britto

## Abstract

Acute pulmonary Exacerbations (AE) are episodes of clinical worsening in cystic fibrosis (CF), often precipitated by infection. Timely detection is critical to minimize the morbidity and lung function decline associated with acute inflammation during AE. We previously demonstrated that the airway protein Short Palate Lung Nasal epithelium Clone 1 (SPLUNC1) is regulated by inflammatory signals. Here, we investigated the use of SPLUNC1 fluctuations to diagnose and predict AE in CF.

We enrolled adult CF subjects from two independent cohorts to measure AE markers of inflammation in sputum and recorded clinical outcomes for a 1-year follow-up period.

SPLUNC1 levels were high in healthy control sputum *(n=9, 10.7μg/mL)*, and significantly decreased in CF subjects without AE *(n=30, 5.7μg/mL, p=0.016)*. SPLUNC1 levels were 71.9% lower during AE *(n=14, 1.6μg/mL, p=0.0034)* regardless of age, sex, CF-causing mutation, or microbiology findings. Cytokines Il–1β and TNFα were also increased in AE,whereas lung function did not consistently decrease. Stable CF subjects with lower SPLUNC1 levels were much more likely to have an AE at 60 days *(Hazard Ratio: 11.49, Standard Error: 0.83, p=0.0033)*. Low-SPLUNC1 stable subjects remained at higher AE risk even one year after sputum collection *(Hazard Ratio: 3.21, Standard Error: 0.47,p=0.0125)*. SPLUNC1 was transcriptionally downregulated by inflammatory cytokines and degraded by proteases increased in sputum during AE.

Our findings suggest that low sputum SPLUNC1 levels could detect subjects at increased risk of AE in order to guide early therapeutic interventions in CF.

**TAKE-HOME MESSAGE:** Sputum concentrations of the secreted airway protein SPLUNC1 decrease during CF exacerbations. Lower SPLUNC1 levels in stable subjects portend a significantly increased risk of exacerbation and could inform therapeutic interventions.

**PLAIN LANGUAGE SUMMARY:** SPLUNC1 is an abundant host defense protein found in the respiratory tract that decreases with inflammation. Individuals with cystic fibrosis experiencing clinical worsening (exacerbation) have much lower levels of SPLUNC1 in their sputum. In stable cystic fibrosis patients, lower levels of SPLUNC1 may predict an upcoming respiratory illness. Therefore, SPLUNC1 may serve as a tool for early diagnosis and treatment of cystic fibrosis exacerbations.

## BACKGROUND

Cystic fibrosis (CF) is one of the most common life-shortening genetic diseases in the United States[1–3]. The primary cause of mortality in CF is lung disease, characterized by persistent airway and lung inflammation leading to lung function decline [4–5]. CF exacerbations (AE) are acute declines in a CF subject’s clinical condition, associated with increased morbidity, healthcare costs, and worsening quality of life [5–9]. AE are associated with increased lung inflammation, manifested as rising concentrations of airway cytokines and proteases that contribute to further lung injury in CF [7‚ 10].

The early diagnosis and treatment of AE is important to minimize morbidity and disease progression in CF. In order to support clinical decision-making, markers of acute inflammation or lung function decline can be used. Decreases in the Forced Expiratory Volume in the first second (FEV1), a measure of pulmonary function, can reflect lung function decline during AE. However, these changes often occur as a late consequence of lung inflammation, limiting its clinical usefulness for early AE detection [11]. Inflammatory cytokines (e.g. IL-6, IL-8, TNFα) have also been linked to AE, but delayed upregulation and limited data on their ability to predict AE limit their clinical usefulness. The development of early non-invasive markers to predict clinical deterioration before FEV1 or cytokine fluctuations would be a significant advancement to detect AE and limit its impact on long-term health in CF[12–14].

In our previous studies, we demonstrated that airway concentrations of host defense protein Short Palate Lung Nasal epithelium Clone 1 (SPLUNC1, also known as BPI-fold containing family member A1, BPIFA1), are closely regulated by microbial and inflammatory signals, as well as proteases [15– 18]. SPLUNC1 is abundant in human respiratory secretions and downregulated within hours by inflammation, suggesting that it could be an early marker of airway irritant exposures or acquisition of pathogens likely to induce clinical deterioration [19].

Here, we hypothesized that SPLUNC1 concentrations in sputum would decrease sharply during AE, and that lower levels of SPLUNC1 would be associated with adverse CF clinical outcomes. We measured SPLUNC1 fluctuations during AE and performed experiments to define the mechanisms that underlie SPLUNC1 regulation. Our findings provide evidence for the use of SPLUNC1 as a marker and predictor of AE that can inform clinical decision-making in CF care.

## MATERIALS AND METHODS

### Detailed methods in Supplemental Methods Section

#### Study Design

This was a two-center, prospective, longitudinal study of CF subjects during periods of clinical stability and AE. All patients received standard-of-care therapy, and cystic fibrosis transmembrane conductance regulator (CFTR) modulators when they became available. The primary objective of this study was to define an association between AE and sputum levels of SPLUNC1. Each subject provided a sputum sample and underwent spirometry within 24 hours of sample collection. Subjects were followed at quarterly outpatient clinic visits, or sooner when indicated, for up to one year (*Supplemental figure 1*). Clinical information, expectorated sputum, and pulmonary function testing data were collected at each clinic visit.

#### Definition of CF Exacerbation

CF AE were defined as the emergence of 4 of 12 signs or respiratory symptoms, prompting changes in therapy and initiation of antibiotics (modified from Fuchs’ criteria [12]). These criteria included: change in sinus congestion, sputum, or hemoptysis; increased cough, dyspnea, malaise, fatigue or lethargy; fever; hyporexia or weight loss; change in chest physical exam; or FEV1 decrease >10% from a previous value [21]. Individuals not meeting AE criteria were characterized as “CF Stable”.

#### Cohort Characteristics

Discovery cohort: 44 adults with confirmed CF diagnosis from the Yale Adult CF Program were recruited during a) scheduled routine visits, b) unscheduled visits in which they reported AE symptoms, and c) on the first day of admission to the hospital for AE treatment. The recruitment period extended from 2014-2016. We organized study subjects in two groups: 1) Stable CF subjects (CF Stable): No new respiratory symptoms, presenting to clinic for scheduled follow up and, 2) AE subjects (AE): Diagnosed with AE (*Table 1*). We also recruited 10 healthy controls (HC) to undergo sputum induction according to published protocols [22]. The study was approved by the Yale University Institutional Review Board and informed consent was obtained from each subject.

**Table 1:** Yale Cohort – Demographics. Demographic characteristics of the Yale Adult CF Program cohort (Discovery Cohort). *HC: Healthy controls; CF Stable: CF subjects without exacerbation; AE: Subjects with active exacerbation*.

| *Number of Patients (n)* | *HC (9)* | *CF Stable (30)* | *AE (14)* |
| --- | --- | --- | --- |
| Age |  |  |  |
| Age (Mean) | 33.5 | 41.1 | 32.1 |
| Age (STD) | 10.7 | 17.0 | 6.4 |
| Age (Range) | 27-45 | 20-79 | 23-43 |
| Sex |  |  |  |
| Female (n) | 1 | 17 | 8 |
| Female (%) | 11.1 | 56.7 | 57.1 |
| Male (n) | 8 | 13 | 6 |
| Male (%) | 88.9 | 43.3 | 42.9 |
| Mutation Background |  |  |  |
| *F508del/F508del* (n) | NA | 11 | 8 |
| *F508del/F508del* (%) | NA | 36.7 | 57.1 |
| *F508del/other* (n) | NA | 11 | 4 |
| *F508del/other* (%) | NA | 36.7 | 28.6 |
| Other mutations (n) | NA | 8 | 2 |
| Other mutations (%) | NA | 26.7 | 14.3 |
| FEV<sub>1</sub> (L) |  |  |  |
| FEV<sub>1</sub> (Mean) | NA | 2.2 | 1.9 |
| FEV<sub>1</sub> (STD) | NA | 0.7 | 0.7 |
| FEV<sub>1</sub> (Range) | NA | 0.57-3.3 | 0.55-2.98 |
| FEV<sub>1</sub> (%) |  |  |  |
| FEV<sub>1</sub> (Mean) | NA | 67.5 | 56.1 |
| FEV<sub>1</sub> (STD) | NA | 24.3 | 24.3 |
| FEV<sub>1</sub> (Range) | NA | 12-121 | 12-89 |
| BMI (Kg/m<sup>2</sup>) |  |  |  |
| BMI (Mean) | NA | 24.4 | 22.7 |
| BMI (STD) | NA | 4.1 | 4 |
| BMI (Range) | NA | 17.6-35.8 | 17.6-35.8 |
| Exacerbations per year (AE/y) |  |  |  |
| AE/y (Mean) | NA | 1.9 | 3.7 |
| AE/y (Range) | NA | 0-10 | 1-10 |
| CF Comorbidities |  |  |  |
| PI (n) | NA | 25 | 14 |
| PI (%) | NA | 83.3 | 100 |
| CFRD (n) | NA | 11 | 9 |
| CFRD (%) | NA | 36.7 | 64.3 |
| Microbiology |  |  |  |
| PA Colonization (n) | NA | 12 | 9 |
| PA Colonization (%) | NA | 40 | 64.3 |
| CFTR Modulators |  |  |  |
| Ivacaftor (n) | NA | 2 | 0 |
| Ivacaftor (%) | NA | 6.7 | 0 |
| Ivacaftor/Lumacaftor (n) | NA | 6 | 7 |
| Ivacaftor/Lumacaftor (%) | NA | 20 | 50 |

FOOTER: Table 1: Demographic characteristics of the Yale Adult CF Program cohort (Discovery Cohort). *HC: Healthy controls; CF Stable: CF subjects without exacerbation; AE: Subjects with active exacerbation.*

Validation cohort: 35 adult and pediatric subjects with a confirmed CF diagnosis, previously enrolled in a sputum study of AE at the University of Minnesota (UMN)were included. This was a prospective study of patients hospitalized for AE treatment [12]. All patients received standard-of-care therapy and each subject provided sputum samples and performed pulmonary function tests within 72 hours of intravenous antibiotic initiation (Table 2)[23].

**Table 2:** University of Minnesota Cohort – Demographics Demographic characteristics of the University of Minnesota CF Centercohort (Validation Cohort). *CF Stable: CF subjects without exacerbation; AE: Subject with active exacerbation*.

| *Age* | *CF Stable (11)* | *AE (24)* |
| --- | --- | --- |
| Age (Mean) | 27.1 | 30.0 |
| Age (STD) | 7.7 | 10.5 |
| Age (Range) | 14-40 | 13-57 |
| *Sex* |  |  |
| Female (n) | 7 | 12 |
| Female (%) | 63.6 | 50 |
| Male (n) | 4 | 12 |
| Male (%) | 36.4 | 50 |
| *Mutation Background* |  |  |
| *F508del/F508del* (n) | 6 | 14 |
| *F508del/F508del* (%) | 54.5 | 58.3 |
| *F508del/other* (n) | 5 | 9 |
| *F508del/other* (%) | 45.5 | 37.5 |
| Other mutations (n) | 1 | 0 |
| Other mutations (%) | 9.1 | 0 |
| FEV<sub>1</sub> (L) |  |  |
| FEV<sub>1</sub> (Mean) | 2.4 | 1.7 |
| FEV<sub>1</sub> (STD) | 0.6 | 0.6 |
| FEV<sub>1</sub> (Range) | 1.39-3.85 | 0.70-2.80 |
| FEV<sub>1</sub> (%) |  |  |
| FEV<sub>1</sub> (Mean) | 67.3 | 45.3 |
| FEV<sub>1</sub> (STD) | 12.1 | 14.1 |
| FEV<sub>1</sub> (Range) | 45.5-86.5 | 26-74 |
| BMI |  |  |
| BMI (Mean) | 21.8 | 21.3 |
| BMI (STD) | 2.1 | 3.1 |
| BMI (Range) | 18.1-24.6 | 13.1-25.8 |
| Exacerbations per year (AE/y) |  |  |
| AE/y (Mean) | 4.1 | 4.3 |
| AE/y (Range) | 1-12 | 1-10 |
| CF Comorbidities |  |  |
| PI (n) | 11 | 23 |
| PI (%) | 100 | 95.8 |
| CFRD (n) | 6 | 9 |
| CFRD (%) | 54.5 | 37.5 |
| Microbiology |  |  |
| PA Colonization (n) | 5 | 9 |
| PA Colonization (%) | 45.5 | 37.5 |
| CFTR Modulators |  |  |
| Ivacaftor (n) | 0 | 0 |
| Ivacaftor (%) | 0 | 0 |
| Ivacaftor/Lumacaftor (n) | 0 | 0 |
| Ivacaftor/Lumacaftor (%) | 0 | 0 |

FOOTER: Table 2: Demographic characteristics of the University of Minnesota CF Center cohort (Validation Cohort). *CF Stable: CF subjects without exacerbation; AE: Subjects with active exacerbation.*

#### Sputum Collection and Processing

CF subjects expectorated sputum spontaneously for airway cultures and provided an additional sample for our study. Induced sputum samples were obtained from HC by sputum induction as previously reported[22, 24]. Sputum was processed as previously reported [20].

#### SPLUNC1 and Cytokine ELISA, Western Blot

We developed and optimized a sputum SPLUNC1 ELISA for this study, which is detailed in the supplemental methods. Human SPLUNC1 recombinant protein was used as a reference (cat# H00051297-P01, Abnova, Taipei, Taiwan). Detection antibody: polyclonal mouse anti-human SPLUNC1 IgG (cat# SAB1401687, MilliporeSigma, Burlington, MA). Secondary antibody: HRP-conjugated anti-mouse IgG (cat# G21040, Invitrogen, Carlsbad, CA). Chromogenic tetramethylbenzidine substrate was applied (KPL, Gaithersburg, MD) and reactions were measured at optical densities of 450 and 550 nm. The assay limits of detection were 1 – 20,000 ng/mL. Mean intra-assay variability: 5.18% (STDEV 1.28%), Inter-assay variability: 18.44% (STDEV 12.95%). For cytokine detection we used multiplexed cytokine ELISA assays (Mesoscale Diagnostics. Rockville, MD, coefficient of variability 25%). Western blots were performed as previously reported, using human neutrophil elastase (NE, hELA2) mouse monoclonal anti-hELA2 IgG (cat# MAB-91671-100, R&D systems) and mouse polyclonal anti-human SPLUNC1 IgG [17]. Densitometry was determined using ImageJ software.

#### Sputum NE Activity and SPLUNC1 Degradation Assays

NE activity was determined using the 7-amino-4-methylcoumarin (MCA) assay (#MAA-3133; Peptides International, Louisville, KY), as previously described [17]. For degradation assays, recombinant human SPLUNC1 was incubated with recombinant human NE (rhNE, R&D systems cat#9167-SE-020) or Pseudomonas aeruginosa elastase (LasB, a gift from Dr. Karen Agaronyan, Yale) at a concentration of 5 μg/ml for 24 hours. SPLUNC1 was measured by ELISA.

#### Regulation of Epithelial Cytokine Expression

Mouse tracheal epithelial cells (mTECs) were isolated from C57BL/6 mice and cultured at air-liquid interface (ALI) as previously described [15]. mTECs were treated with recombinant murine IL-1β (cat# 211–11b, Peprotech, Rocky Hill, NJ) or TNF-α (cat# 315-01A, Peprotech), at 10 ng/mL for 24 hours. NCI-H292 human airway epithelial cells (shown to abundantly express SPLUNC1 and to be robustly regulated by inflammatory cytokines [15]), were treated with recombinant human IL-1β (cat# PHC0811, Gibco, Gaithersburg, MD) or TNF-α (cat# 210-TA-005, R&D, Minneapolis, MN) at 10 ng/ml. Cellular mRNA was extracted for qPCR, and SPLUNC1 qPCR assays were performed to quantify *SPLUNC1* transcriptional regulation as described[15].

#### Statistical Analysis

Descriptive statistics were calculated for the entire subject population. Pearson correlations, or Spearman correlations for variables that were not normally distributed, were calculated between SPLUNC1 and clinical parameters. In order to select the optimal threshold of SPLUNC1 and cytokine concentrations that identify a subject group at higher AE risk, we calculated receiver-operating curves (ROC) based on the distribution of SPLUNC1, IL-1β, TNFα, G-CSF, IL-6, and IL-8 levels in the discovery cohort (*Supplemental Figure 3*). Using this threshold, we applied statistical modeling (Mantel-Haenszel estimator) to predict AE-free intervals. AE-free interval was defined as the time in days from sputum sampling in a stable subject to the time of the first AE after that visit. Cox proportional hazards model was conducted with clinical parameters as covariates. A backward elimination strategy with a significance level to stay of 95% (a=0.05) was employed to achieve a parsimonious model. All statistical analyses were conducted using SAS 9.4 with a level of significance of 95%(α=0.05).

## RESULTS

### SPLUNC1 is Decreased in the Sputum of Stable CF Subjects

SPLUNC1 levels ranged from 4.41 to 22.24μg/mL in the sputum of healthy controls (HC). In stable CF subjects (CF Stable), SPLUNC1 was significantly decreased, whereas total sputum protein was increased (*Figure 1A, 1B*). To further characterize the inflammatory profile of stable CF subjects, we measured sputum concentrations of inflammatory cytokines previously reported to be increased in CF. Of these, IFNα, IFNYγ, IL1β, IL-8, IL-13, and TNFα were significantly increased in CF (*Figure 1C*). When separated according to CF-causing mutation genotype, there were no differences in SPLUNC1 levels of stable subjects with homozygous, heterozygous, or no F508del genotype (*Supplemental Figure S2*). Furthermore, SPLUNC1 was decreased in CF regardless of the presence of CF-related complications, microbiology findings, and CFTR modulator therapies (Table 1 and 2). These findings indicate that SPLUNC1 is abundant in sputum and decreased in stable CF subjects.

**Figure 1.**
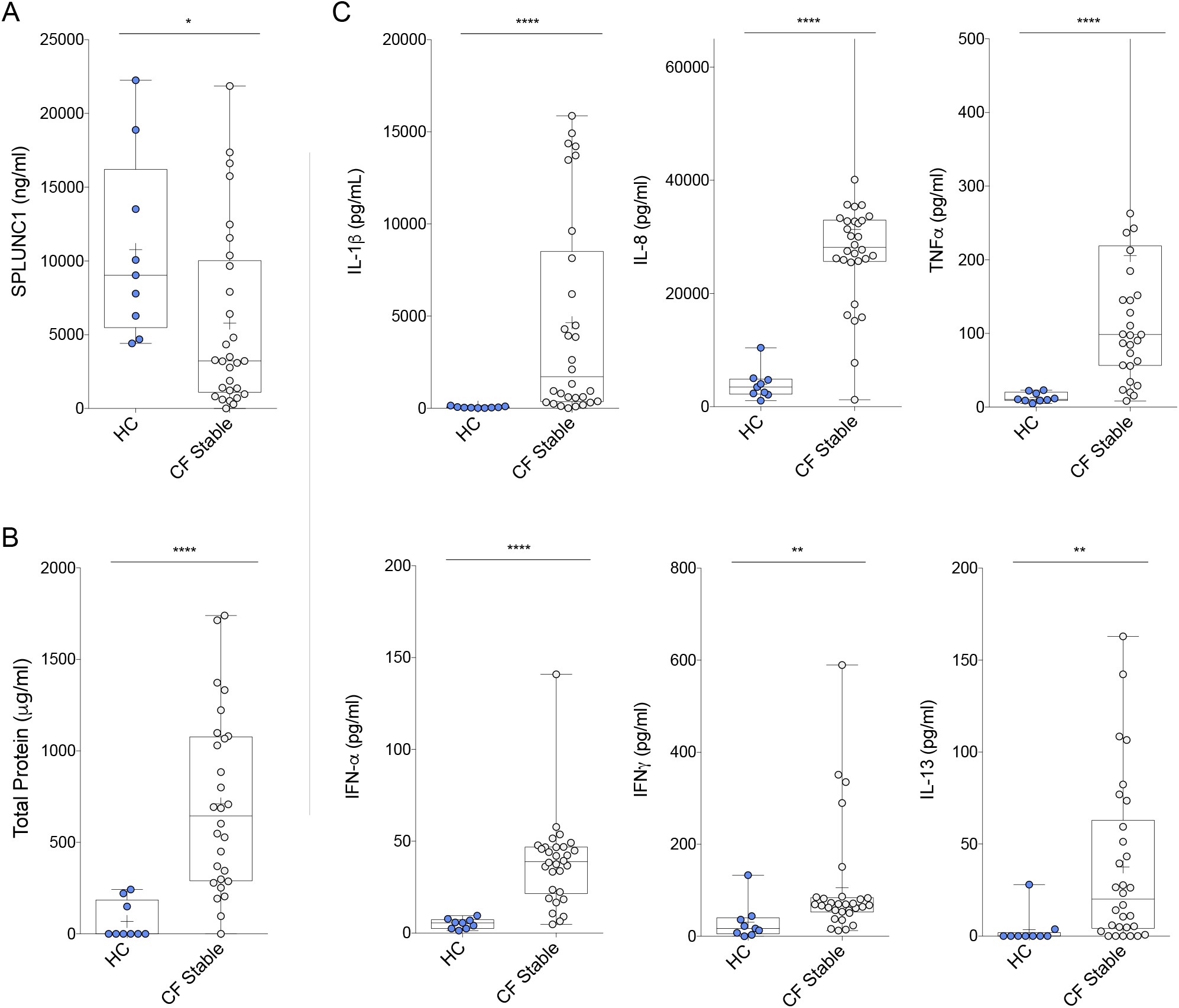
SPLUNC1 is Decreased in the Sputum of Stable CF Subjects. **A)** SPLUNC1 levels (ELISA) in sputum samples from the Yale cohort of adult CF subjects without respiratory symptoms (*CF Stable*) and healthy controls (HC). **B)** Total protein in sputum (BCA assay) from the same subjects. C) Inflammatory cytokine levels(ELISA) in sputum from the same subjects. Additional cytokines tested without significant difference: CXCL10, G-CSF, IFNλ, IL-6, IL-13, MCP1, MIP1α. CF Samples were obtained by voluntary expectoration during clinical assessment, HC samples obtained by sputum induction with nebulized normal saline solution. *+ = Mean; Bar inside box: Median; Whiskers: Minimum/Maximum. Mann-Whitney Test with Bonferroni correction; * = p<0.05; ** = p<0.01; **** = p<0.0001*.

### SPLUNC1 is Decreased During CF Exacerbations

AE are frequently caused by new airway infection and increased airway inflammation. Therefore, we hypothesized that SPLUNC1 levels would decrease during AE. To test this, we measured SPLUNC1 levels in sputum from stable CF and AE subjects. SPLUNC1 decreased sharply during AE in the discovery cohort (Yale University, 71.9% decrease) and in the validation cohort (UMN, 38.6% decrease) (Figure 2). In contrast,FEV1, a marker of lung function widely used to diagnose AE, did not decrease in the Yale AE group, but was significantly decreased in the UMN AE group.

We hypothesized that individuals receiving intravenous antibiotics (IV) for AE would be more severely ill and have more airway inflammation than those treated with oral antibiotics, thus IV-treated subjects would have larger drops in SPLUNC1. To test this, we measured SPLUNC1 in sputum of AE subjects being treated with oral antibiotics as outpatient (AEOP) or IV antibiotics as inpatient (AEIP) in the UMN cohort. In both treatment groups, SPLUNC1 was lower than in stable CF subjects, however there was no difference between AEOP and AEIV levels of SPLUNC1 (Supplemental Figure S4). This indicates that acute drops in sputum SPLUNC1 occur consistently during AE regardless of AE severity.

**Figure 2.**
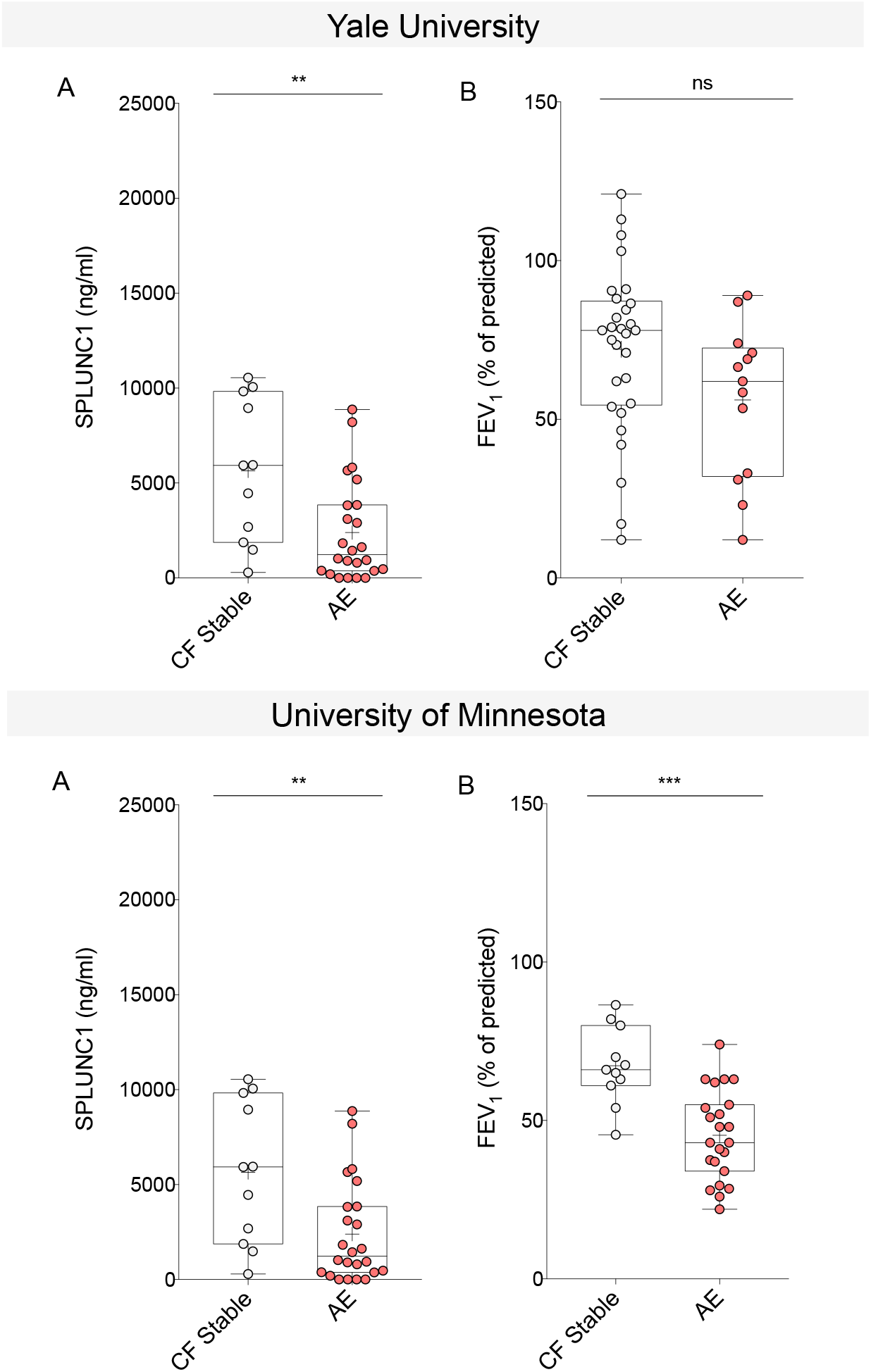
SPLUNC1 is Decreased During Acute CF Exacerbations (AE). Sputum SPLUNC1 and FEV1 from two clinical cohorts including adult (Yale University, n=44) and mixed adult/pediatric (University of Minnesota, n=35) CF subjects. Samples were obtained by voluntary expectoration during clinical assessment, A) SPLUNC1 quantified by ELISA, B) FEV_1_ (Percent of Predicted, %) obtained by spirometry during clinical assessment; *CF Stable: No symptoms of AE, no antibiotictreatment. AE: Acute CF exacerbation, symptoms of AE and ongoing antibiotic therapy; FEV_1_: Forced Expiratory Volume in the first second; + = Mean; Bar inside box: Median; Whiskers: Minimum/Maximum. Mann-Whitney test; ** = p<0.005; *** = p<0.001; ns = not statistically significant*

Next, we sought to define SPLUNC1 fluctuations during AE within the same individuals, relative to their stable-state reference value (subject-specific fluctuations). We compared SPLUNC1 and FEV_1_, (%) in paired samples from the same subjects, collected during stable and AE periods. SPLUNC1 decreased during AE in both the Yale and UMN cohorts (*Figure 3*). In contrast, FEV_1_, only decreased during AE in the UMN cohort. These findings suggest that SPLUNC1 is consistently decreased during AE, while FEV1 decreases during AE vary across cohorts.

**Figure 3.**
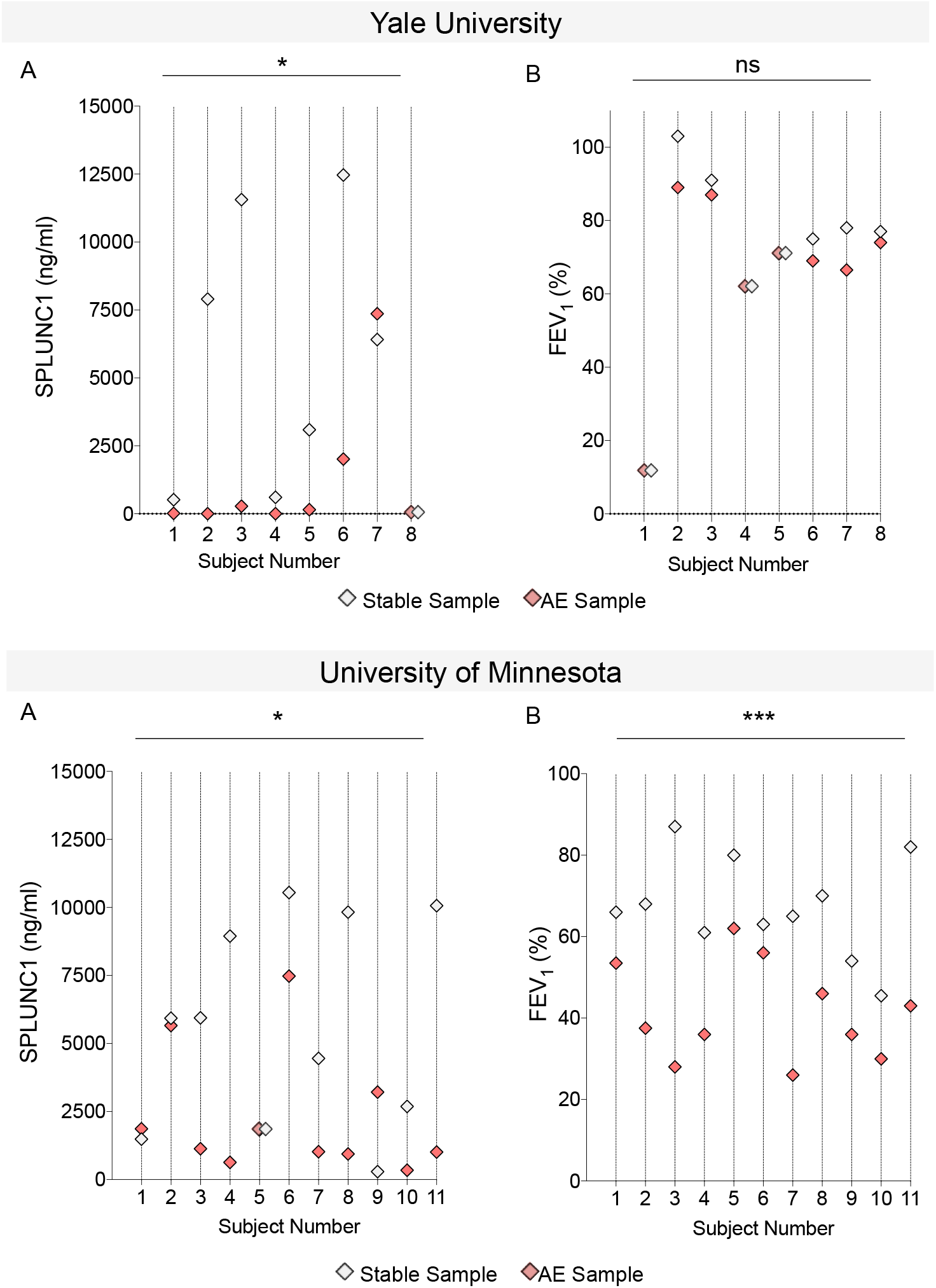
Subject-Specific SPLUNC1 and FEV1 Decreases During AE. **A)** Paired SPLUNC1 levels in sputum samples from the same individual with and without AE (ELISA); **B)** Paired FEV1 measurements from the same individual with and without AE (Percent of Predicted, %) obtained by spirometry during clinical assessment; Samples from two clinical cohorts including adult (Yale University, n=8) and mixed adult/pediatric CF subjects (University of Minnesota, n=11). Each vertical line and number represent a single subject that provided one Stable and one AE sample. *CF Stable (Gray markers): No symptoms of AE, no antibiotic treatment.AE (Red markers): Acute CF Exacerbation, symptoms of AE and ongoing antibiotic therapy; When values were the same, these were represented by two overlapping diamonds along the subject’s line. Wilcoxon matched-pairs signed rank test;* = p<0.05, *** = p = 0.0001, ns=not statistically significan*

### Low SPLUNC1 Levels in Sputum Predict AE-Free Time in Stable CF Subjects

To determine if SPLUNC1 is a predictor of AE-free time, the number of days from sputum collection in stable patients to the next time they were diagnosed with AE, we performed a Mantel-Haenszel survival estimator analysis for AE-free time after separating the groups into high- and low-SPLUNC1 (Detailed methods, *Supplemental Figure 3 5*). In stable CF subjects, the SPLUNC1-low group had a median AE-free time of 43.5 days compared to 150 days in the SPLUNC1-high group. This indicates that high SPLUNC1 levels are associated with more AE-free days.

Next, we performed Cox-proportional Hazards modeling to assess the likelihood of AE when adjusting for demographics, CF-related comorbidities, microbiology, and lung function. SPLUNC1 remained a strong predictor of AE-free time regardless of *F508del* mutation status, microbiology, or severity of FEV_1_, impairment. Importantly, subjects in the SPLUNC1-low group were at significantly increased short- and long-term risk of AE (Hazard ratios: 11.49 at 60 days p=0.003, *Figure 4A*) and 3.21 at 1 year (p=0.013, *Figure 4B*).

**Figure 4.**
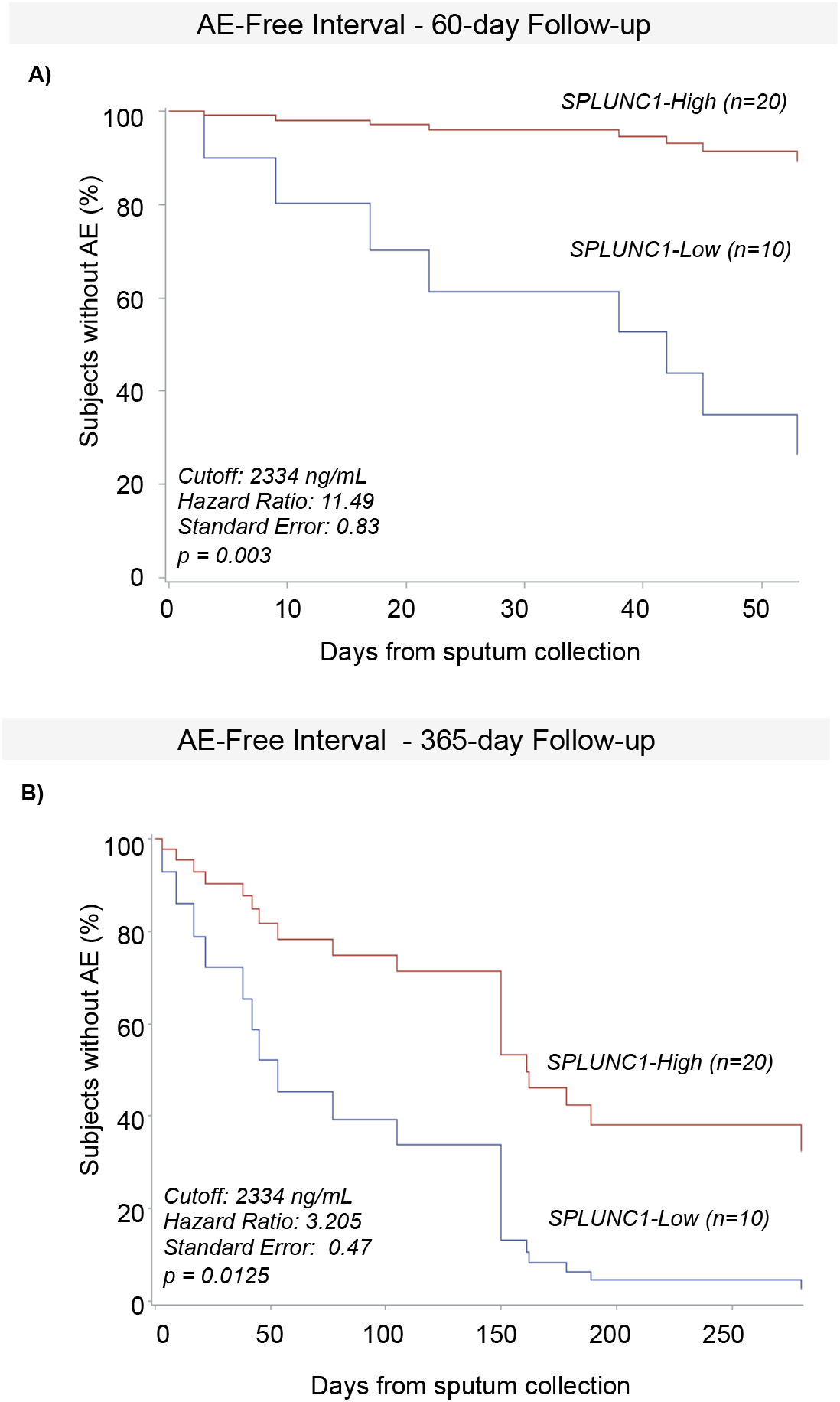
SPLUNC1 Predicts AE-Free Time. **A)** AE-Free time in StableCF subjects separated into SPLUNC1-High and SPLUNC1-Low groups over a 60-day follow up period. SPLUNC1-high and -low groups were defined according to sputum concentration thresholds obtained from ROCs separating CF Stable and AE levels (*Supplemental Figure 3*). AE-Free time was defined as the number of days from sputum collection in Stable subjects until the date of their next AE. **B)** AE-Free time inStable CF subjects separated into SPLUNC1-High and -Low groups over a 365-day follow up period. Cox Proportional Hazards model used to calculate AE-free intervals and adjust for age, sex, BMI, FEV1, number of F508del mutations, presence of CF-related diabetes or pancreatic insufficiency, use of CFTR modulators, and microbiology for *P. aeruginosa, A. xylosoxidans, H. parainfluenzae, Methicillin-sensitive S. aureus, and Methicillin-resistant S. aureus*.

In order to compare SPLUNC1 to previously reported AE markers as predictors of AE, we defined ROC thresholds and AE-free time for G-CSF, IL1-β, IL-6, IL-8, and TNFα(*Supplemental Figure 3*). In a similar multivariate proportional hazards model, cytokine high/low groups based on these markers did not show an increased hazard ratio of AE at 60 days, and only IL-1β was associated with an AE increased risk at 1 year of follow up (Hazard ratio: 3.90, p=0.017, *Supplemental Figure 6*). These findings suggest that SPLUNC1 is a better predictor of AE risk in the short and long term than previously reported sputum markers of AE.

### Human and Bacterial Elastases Found in CF Sputum Degrade SPLUNC1

In order to understand the mechanisms of SPLUNC1 decrease in AE, we first tested the role NE and bacterial elastases increased during AE in SPLUNC1 degradation. We incubated recombinant human SPLUNC1 (rhSPLUNC1) with recombinant human neutrophil elastase (NE) or *Pseudomonas aeruginosa’s* Elastase B (LasB) at increasing concentrations for 3 and 8 hours. NE and Lasb induced a concentration-dependent decrease in full-length SPLUNC1 (*Figure 5A, B*). Next, we quantified NE concentrations in HC and CF sputum during stable and AE periods. NE was increased overall in CF, but it did not increase significantly from stable levels during AE (*Figure 5C*). Finally, to define subject-specific NE and SPLUNC1 changes, we performed Western blot of HC and CF sputum and probed for NE, followed by re-probing for SPLUNC1. SPLUNC1 was decreased in CF in association with increased NE, however, there were no differences in NE or SPLUNC1 between Stable and AE CF sputum (*Figure 5D*). Our data indicate that NE is important for SPLUNC1 degradation in CF sputum but its concentration does not vary between stable and AE states.

**Figure 5.**
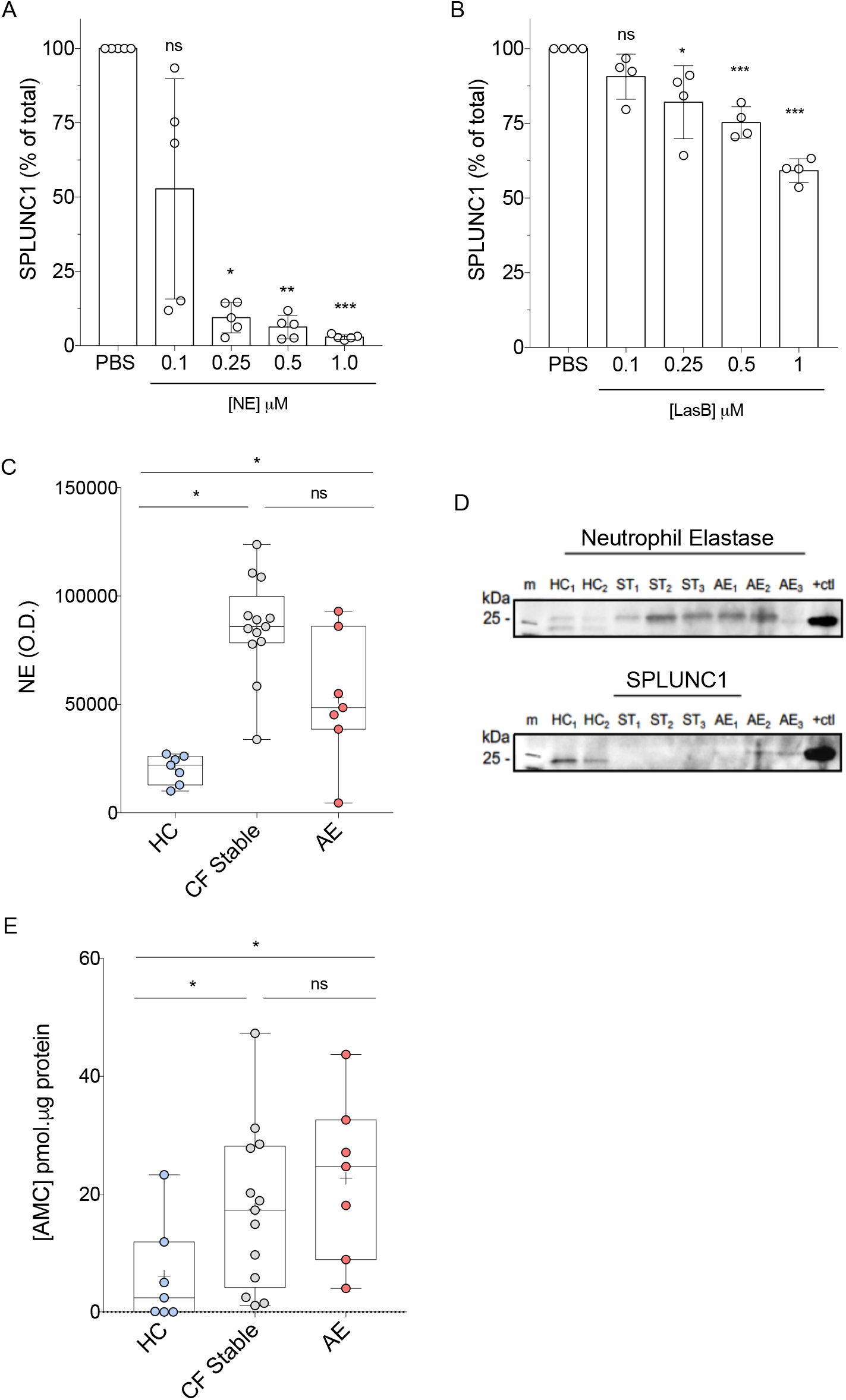
Elastase Concentration and Activity are Increased in CF. **A)** SPLUNC1 densitometry showing degradation by human neutrophil elastase (NE) relative to PBS control, at specified concentrations over 3 hours at 37°C. **B)** SPLUNC1 densitometry showing degradation by Elastase B (LasB) from P. aeruginosa relative to PBS control at specified concentrations over 8 hours at 37°C. **C)** NE densitometry in sputum from healthy controls (HC), Stable CF subjects (CF Stable,and AE subjects (AE) assessed by immunoblot. **D)** Representative blots showing endogenous expression of SPLUNC1 (25 kD) and NE (25–30 kD) in HC and CF sputum samples from the same individual. Membranes were probed for NE prior to stripping and re-probing for SPLUNC1. E) NE Activity in CF sputum: AMC formation from florigenic NE substrate MAA-3133 following 6 h incubation with HC, CF stable, and AE sputum at 37°C. *For experiments in A and B: n = 4–5, 2 individual experiments; Mann-Whitney Test; + = Mean; Bar insidebox: Median; Whiskers: Minimum/Maximum; * p < 0.05; ** = p < 0.01; *** = p < 0.005; ns: not statistically significant; HC: Healthy Control; ST: Stable CF; AE: CF exacerbation; m: marker; +ctl: positive control; OD: optic density*

To determine if NE activity, rather than concentration, increased during AE we measured NE-specific fluorescent cleavage products. When incubated with NE, CF sputum had much higher NE activity than HC sputum; however, there was no difference between stable and AE subjects (Figure 5E).

### SPLUNC1 Expression is Decreased by AE-Associated Cytokines

To further define the AE inflammatory profile of CF subjects, we measured inflammatory cytokines in the sputum of stable and AE subjects. When comparing CF disease states, only IL-1β and TNFα were significantly increased during AE (*Figure 6A*). In order to determine if increased IL-1β and TNFα contributed to decreased SPLUNC1 during AE, we treated primary mouse tracheal epithelial cells (mTEC) and the human airway epithelial cell line NCI-H292 with these cytokines and measured SPLUNC1 mRNA expression. At concentrations encountered in AE sputum, both IL-1β and TNFα decreased *SPLUNC1* expression by airway epithelial cells (*Figure 6B*). Together with our observations from NE and LasB experiments, these findings suggest that during AE, SPLUNC1 is decreased through protein degradation and cytokine-driven transcriptional regulation

**Figure 6.**
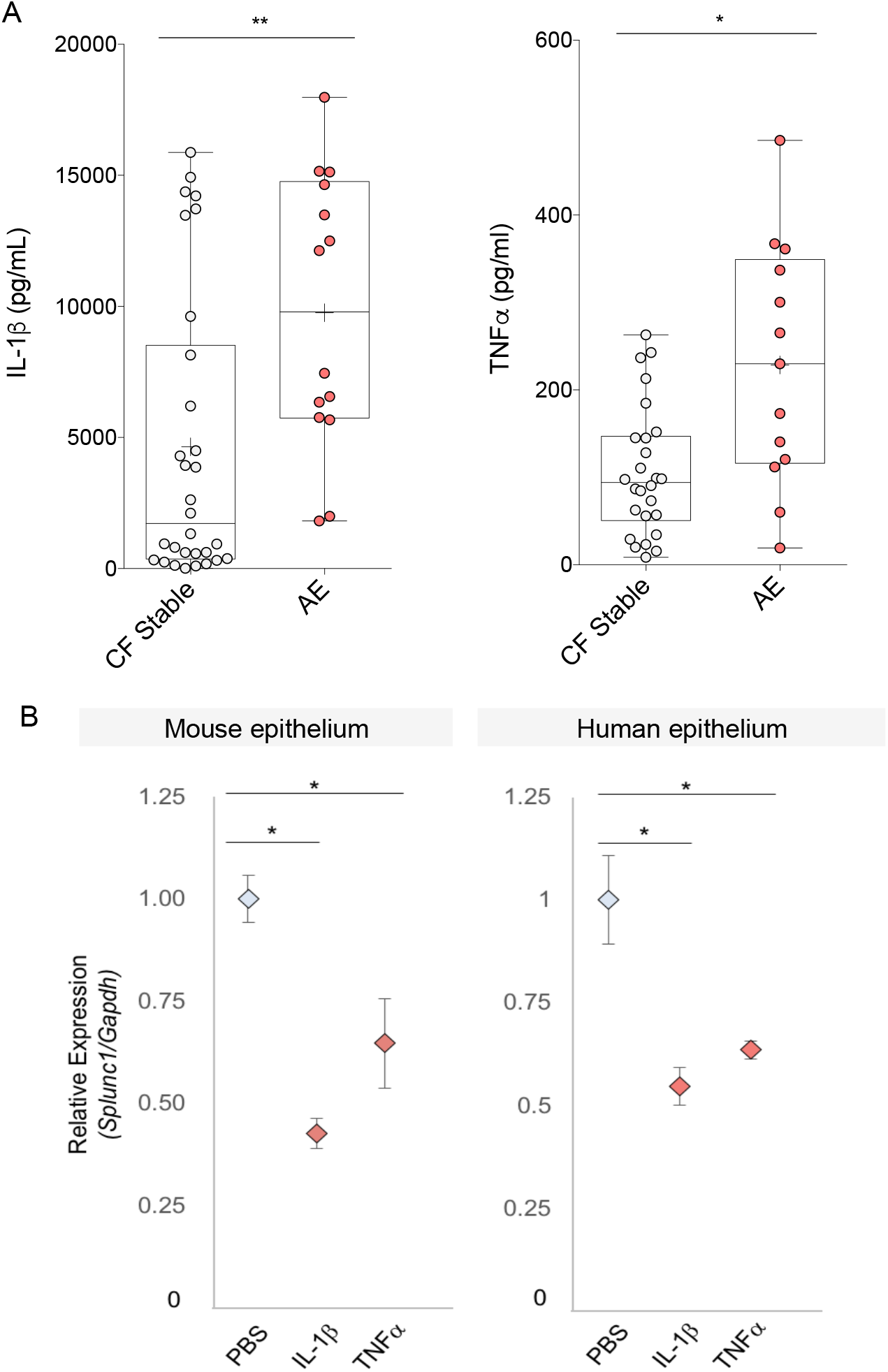
Cytokines Il-1β and TNF-α Increase During AE & Downregulate SPLUNC1 Expression. **A)** Inflammatory cytokine levels in sputum from adult CF subjects uwithout respiratory symptoms (CF Stable) and with acute CF Exacerbation (AE). *Additional cytokines tested without significant difference: CXCL10, G-CSF, IFNα2, IFNλ, IFNλ, IL-6, IL-8, IL-13, MCP1, MIP1a. Mann-Whitney Test with Bonferroni corection*. **B)** Relative SPLUNC1 mRNA expression in mouse tracheal epithelial cells (Mouse epithelium) grown at air-liquid interface and in the NCI-H292 human airway epithelial cell line (Human epithelium) treated with recombinant TNFα and Il-1β (10 ng/mL) for 24hours *(n=4–5/condition, 2 experiments, 2-way ANOVA); + = Mean; Bar inside box: Median; Whiskers: Minimum/Maximum*; mRNA expression quantified by qPCR. * = *p<0.05; ** = p<0.01*

## DISCUSSION

AEs contribute to accelerated lung function decline and increased morbidity and mortality in CF[5–9, 25,26]. Early AE detection is crucial in CF care, however noninvasive inflammatory AE biomarkers are not routinely used in clinical practice. Here, we describe a novel role for SPLUNC1 as a marker and predictor of AE in CF.

Sputum SPLUNC1 is lower in CF overall and further decreased during AE. Importantly, we show that stable subjects with low SPLUNC1 levels have a significantly increased likelihood of AE in the short and long term. These findings suggest that SPLUNC1 levels could inform the diagnosis and clinical management of AE.

The relationship between airway infection, lung inflammation, and lung function decline in CF has been studied in detail [27–29]. In the context of acute airway inflammation, SPLUNC1 decreased within three hours after exposure to airway proteases, and its expression decreased within 24 hours in response to inflammatory cytokines. Pathogens could also decrease SPLUNC1 during AE, as shown here with LasB, an extracellular PA protease detected in CF sputum [30]. Thus, synergistic degradation by inflammatory cells and bacteria, and transcriptional downregulation by IL-1β and TNFα decrease SPLUNC1 as a terminal result of immune activation during AE.

We observed subject-specific SPLUNC1 decreases during AE. This supports measuring baseline levels longitudinally in CF to define a SPLUNC1 baseline before and after development of new respiratory symptoms. This longitudinal baseline could account for daily SPLUNC1 fluctuations related to environmental exposures. Daily SPLUNC1 variations can be differentiated from AE through their dose-effect regulation. For example, basal pathogen and cytokine signals modestly suppress SPLUNC1 levels at baseline. In mouse models lacking these suppressive signals (i.e. IFNγ receptor-deficient, Toll-like receptor 4-deficient, and TLR adaptor molecule MyD88-deficient mice) SPLUNC1 levels are dramatically increased[15]. While daily changes in inflammatory signals or pathogen exposures may cause small fluctuations, SPLUNC1 changes during AE occur rapidly and in much greater magnitude (*Figure 2, 3*). Thus, even in the absence of day-to-day measurements, SPLUNC1 can be a helpful tool to diagnose early or imminent AE.

Low SPLUNC1 levels in stable subjects may also contribute to pathogenesis. SPLUNC1 has host protective functions relevant to CF, including regulation of airway surface liquid, antimicrobial properties, and immunomodulatory effects [31–37]. For example, SPLUNC1 gene polymorphisms causing lower SPLUNC1 levels were recently linked to severe lung function impairments in CF [38]. Although we did not focus on understanding how low SPLUNC1 levels increase AE risk, we interpret that SPLUNC1 cleavage by proteases at functional sites may disrupt its host defense functions[39, 40].

Our study has several limitations. First, we use of a cross-sectional study to develop longitudinal predictions, however this approach enabled us to predict AE in the short and long term with statistically significant findings. Second, our study has a relatively small size, however, our findings show that it was adequately powered to demonstrate differences in key observations that were confirmed on a validation cohort. Finally, although we did not detect differences in stable SPLUNC1 levels between subjects of different CFTR genotypes, our AE study population did have a higher prevalence of advanced lung disease and CF-causing mutations linked to severe disease. We addressed this by using multivariate models to demonstrate that the predictive ability of SPLUNC1 is not affected by these variables. Larger prospective studies are needed to replicate our findings in regards to *non-F508del* genotype cohorts, presence of specific CF comorbidities, and the impact of combined CFTR modulator therapies.

Our findings suggest that SPLUNC1 levels could inform clinical decision-making early in the development of AE, when airway inflammation has not yet translated into detectable lung function decline, overcoming a key limitation of FEV_1_; measurements for the diagnosis of AE. As novel CF therapies become available, noninvasive and airway-relevant biomarkers like SPLUNC1 may be useful for clinical trial candidate recruitment and longitudinal assessments of lung disease control that inform the care of our CF population.

**Supplemental Figure 1.**
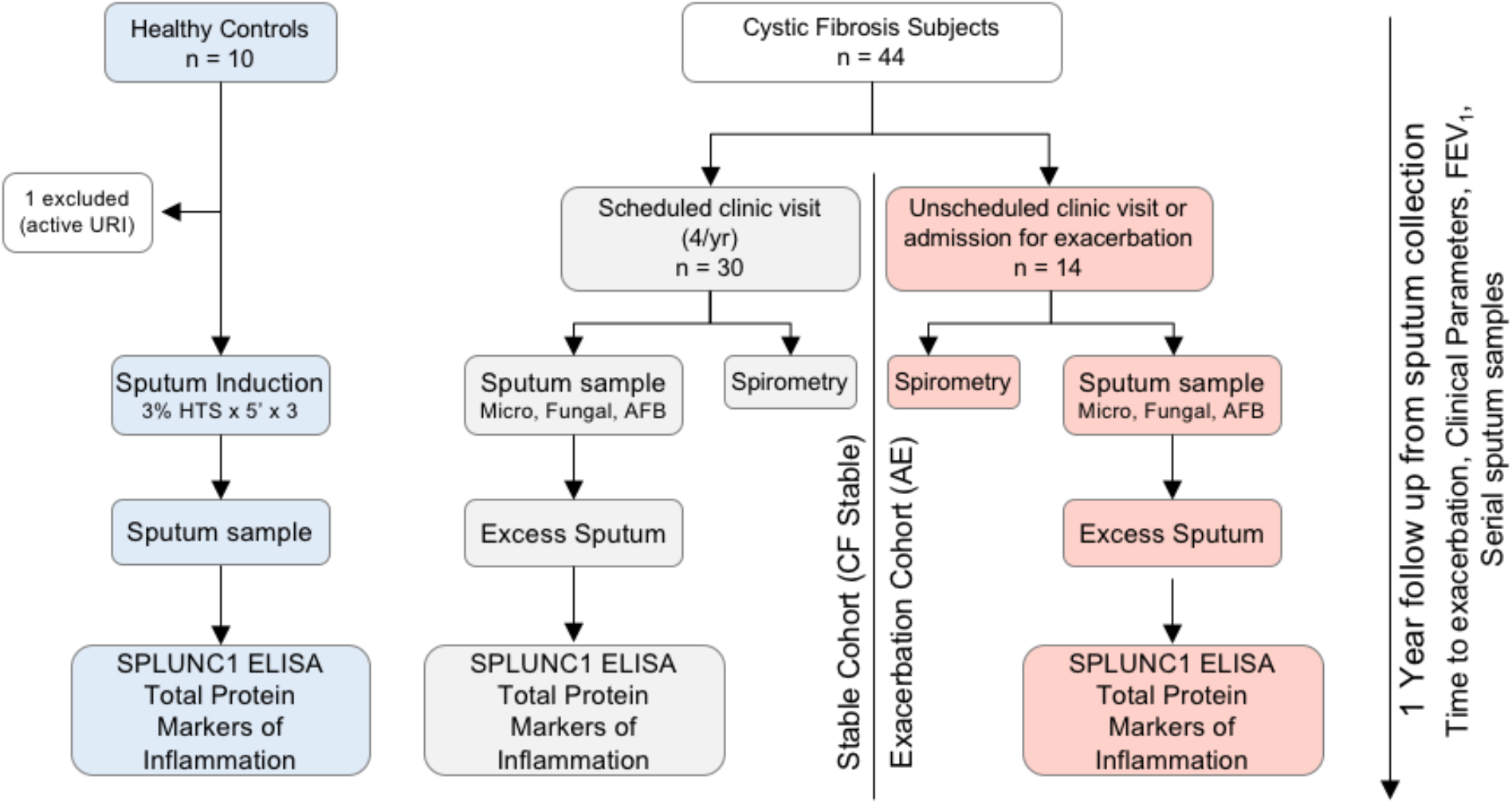
Study design (Yale discovery cohort). Forty-four adult subjects with a confirmed diagnosis of CF were identified from the Yale Adult CF Program to participate in this study. These patients were recruited during their scheduled routine visits, unscheduled “sick” visits in which they reported new respiratory symptoms, and on their first day of admission to the hospital for treatment of a pulmonary exacerbation (AE). Our recruitment period extended from 2014-2016. We organized study subjects into two groups: 1) Stable CF subjects (CF Stable): Individuals without new respiratory symptoms, who presented to clinic for their scheduled quarterly follow up and, 2) Subjects having an AE: Individuals with new respiratory symptoms, clinically diagnosed with AE that were prescribed treatment for AE during scheduled visit, unscheduled sick visit, or first day of hospital admission for AE. All CF subjects provided spontaneously expectorated sputum samples, sputum microbiology samples, and pulmonary function tests. Healthy controls underwent sputum induction. All subjects were followed for the development of AE for one year counted form the date of sputum collection.

**Supplemental Figure 2.**
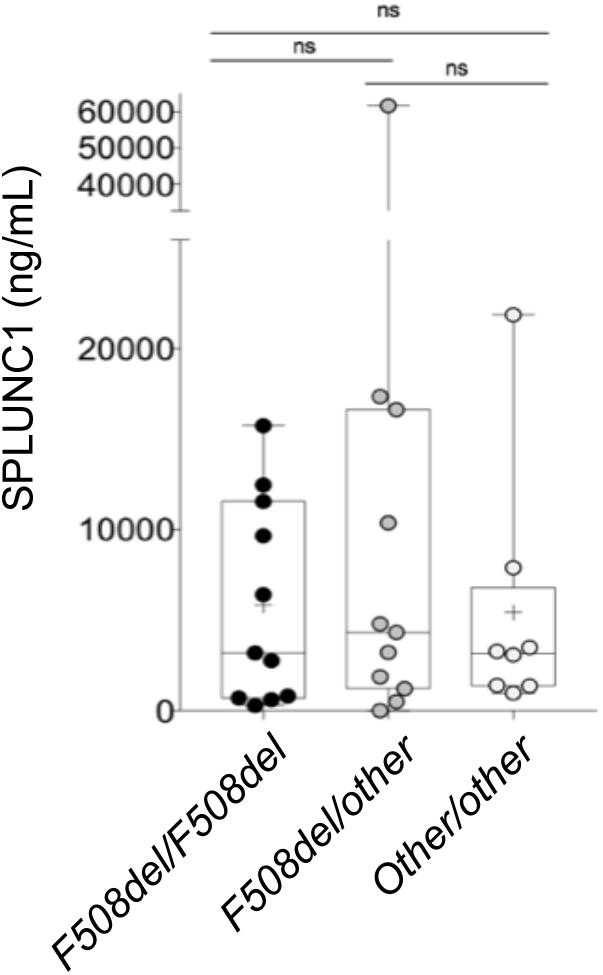
SPLUNC1 does not change according to *F508del* genotype. SPLUNC1 levels (ELISA) in sputum samples from the Yale cohort of adult CF subjects without respiratory symptoms (*CF Stable*) organized by the presence of one, two, or no *F508del* mutations. Sputum Samples were obtained by voluntary expectoration during clinical assessment, + = *Mean; Bar inside box: Median; Whiskers: Minimum/Maximum. Mann-Whitney Test with Bonferroni correction*; ns = *not statistically significant.*

**Supplemental Figure 3.**
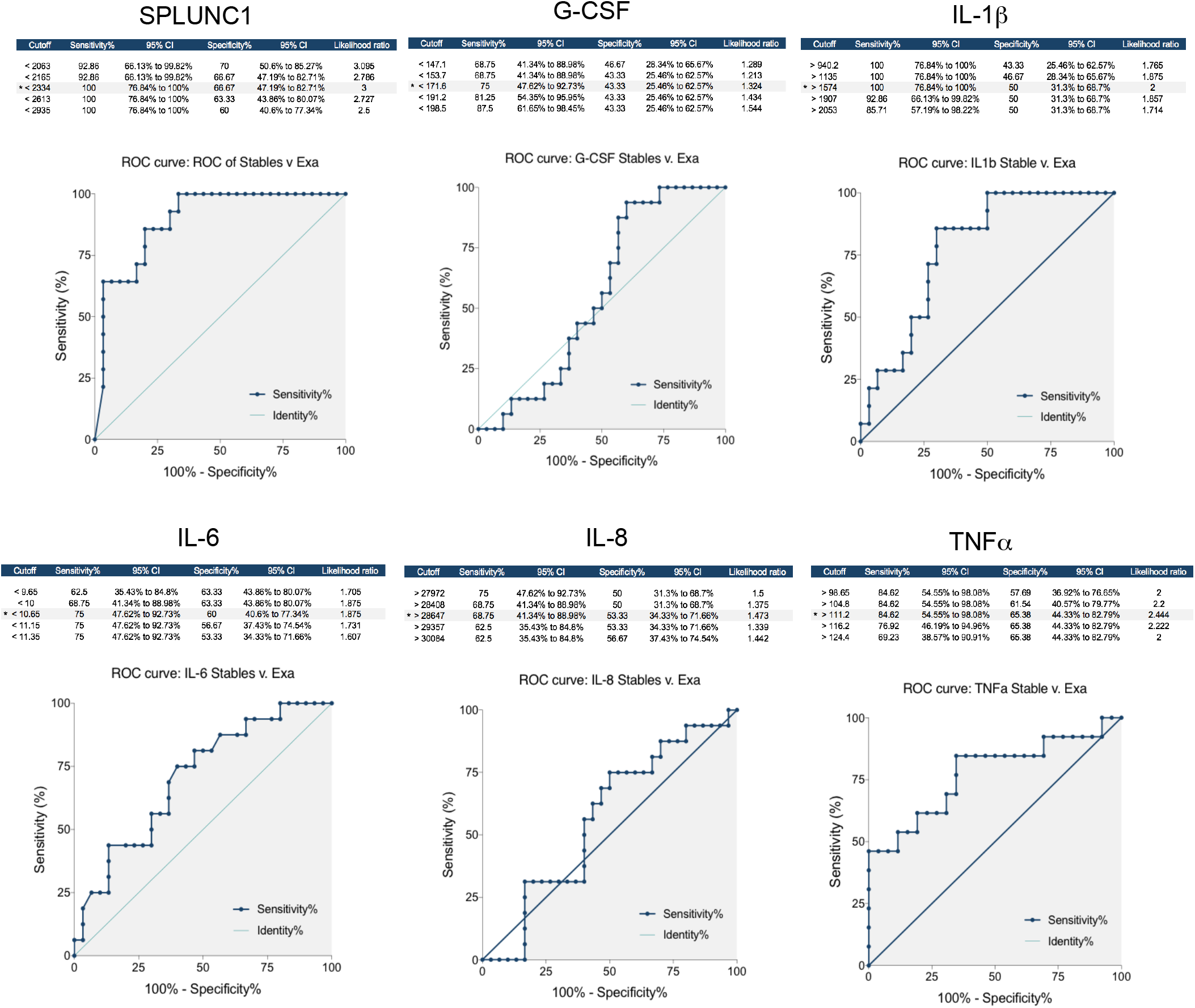
Receiver Operator Curve Development for defining SPLUNC1, IL-1β, TNFα, GCSF, IL-6, and IL-8 High and Low categories. Cutoff values (*) were selected to provide maximum sensitivity with the highest specificity possible (highlighted in gray).

**Supplemental Figure 4.**
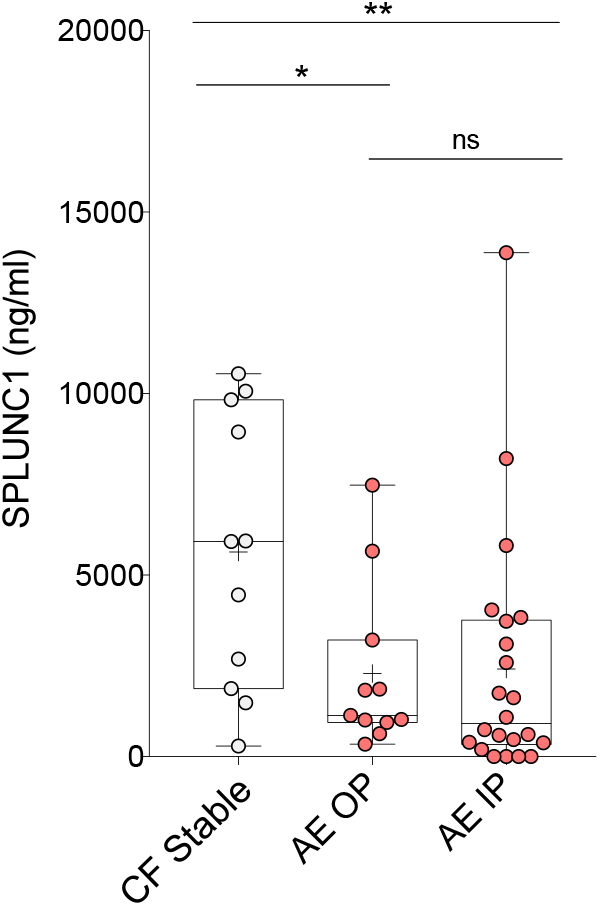
SPLUNC1 is Decreased During CF Exacerbations Requiring Outpatient or Inpatient Antibiotic Treatment. SPLUNC1 levels in sputum samples from a clinical cohort including adult and pediatric CF subjects (University of Minnesota). Samples were obtained by voluntary expectoration during clinical assessment, SPLUNC1 quantified by ELISA. *CF Stable:* No symptoms of AE, no antibiotic treatment. *AE Outpatient (AE OP):* Clinical symptoms consistent with exacerbation, treated with oral antibiotics at the time of sputum collection. *AE Inpatient (AE IP):* Admitted for inpatient antibiotic course, sample collected during first day of treatment. *+ = Mean; Bar inside box: Median; Whiskers: Minimum/Maximum; Mann-Whitney test*; * = *p<0.05*, ** = *p<0.01, ns = not statistically significant*.

**Supplemental Figure 5.**
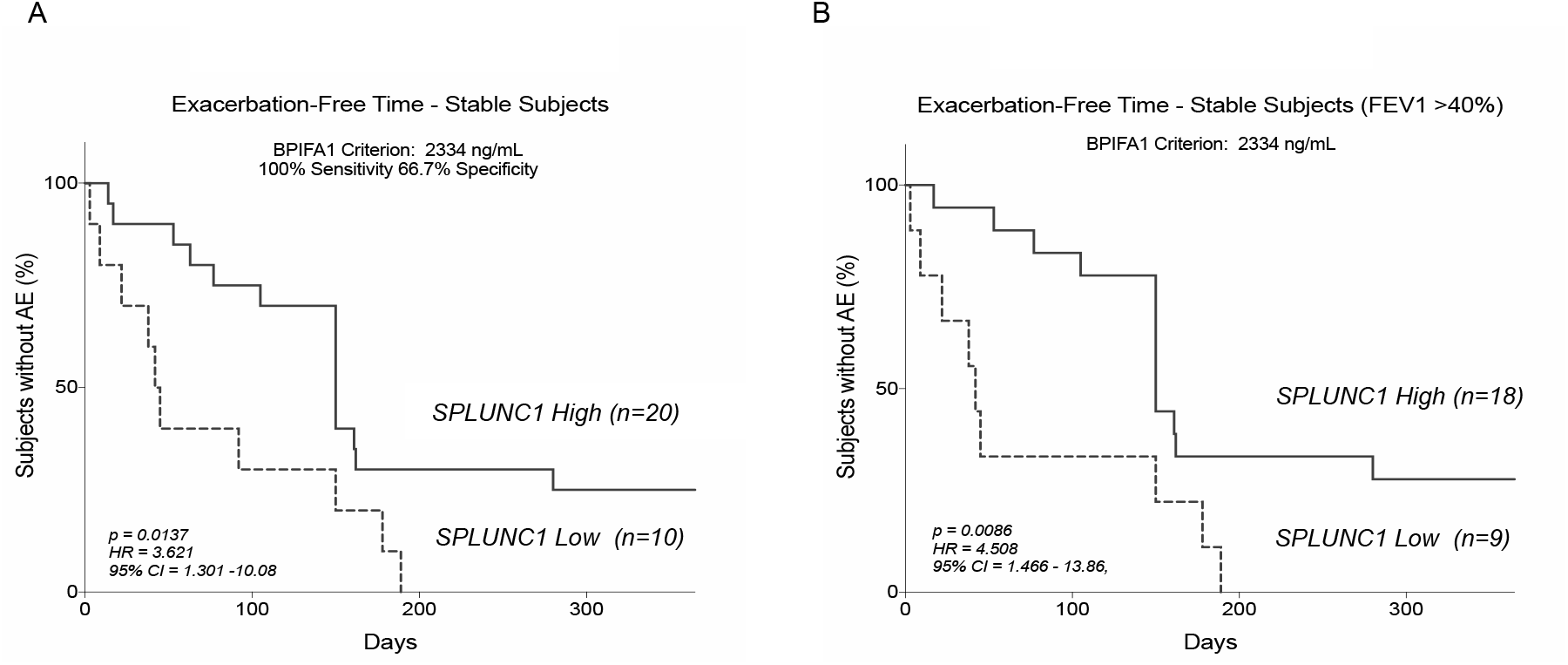
SPLUNC1 Predicts AE-Free Time (unadjusted survival model). **A)** Stable CF subjects were separated into SPLUNC1-high and SPLUNC1-Low groups according to a SPLUNC1 threshold of 2334 ng/mL. Time to AE in days was measured over 365 days from the date of sputum collection in all subjects. **B)** Stable subjects with an FEV_1_ >40% of predicted were separated into SPLUNC1-high and SPLUNC1-low cohorts as above. Time to exacerbation was measured for up to one year from the date of sputum collection. Mantel-Haenszel estimator was used to calculate exacerbation-free interval in each group. Values are not adjusted for clinical variables. Adjusted values from a Cox proportional hazards model are presented in Figure 6. *AE: CF exacerbation, HR: Hazard ratio, CI: Confidence interval*.

**Supplemental Figure 6.**
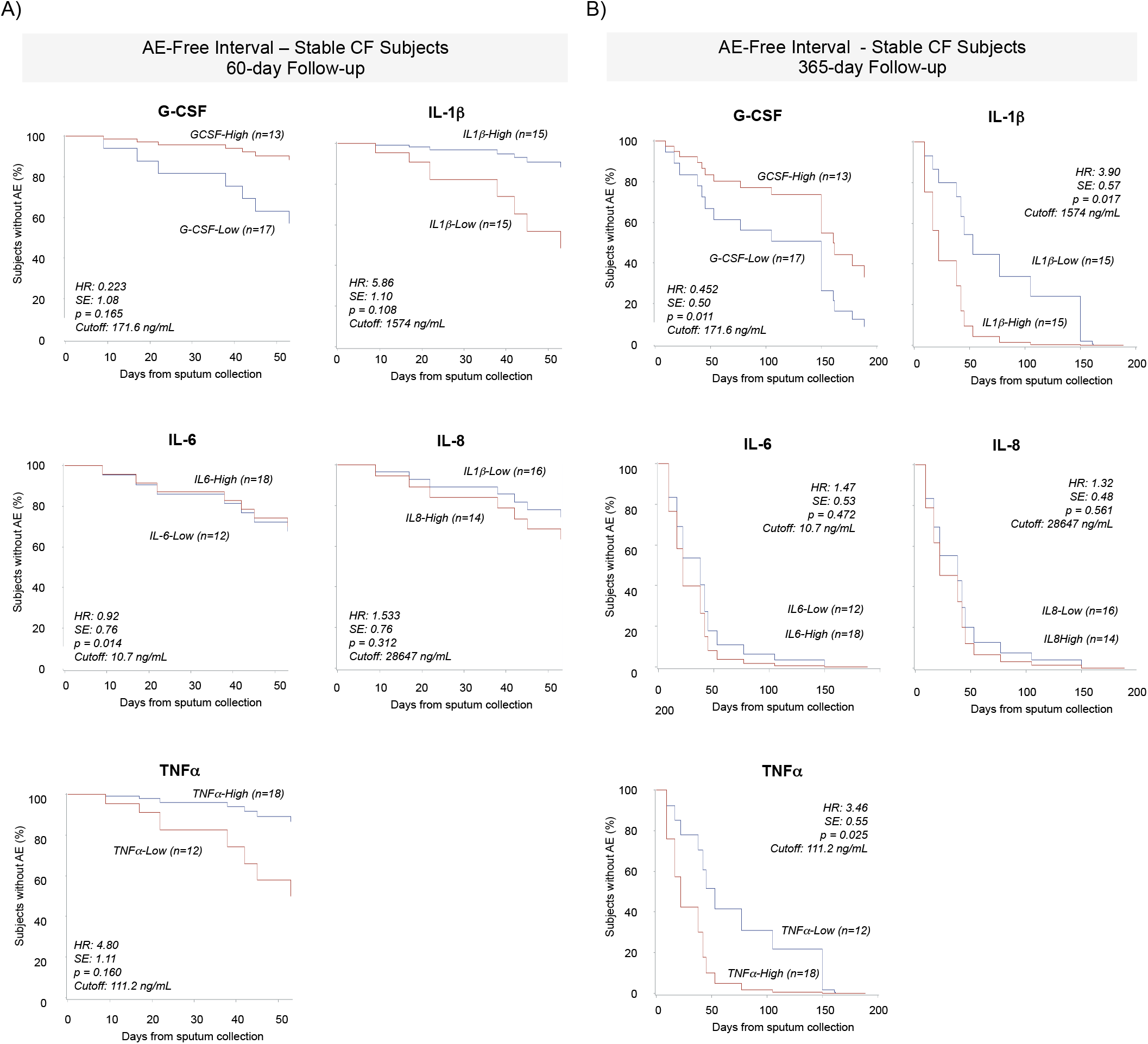
Sputum Cytokines Do Not Predict Short Term AE-Free Time. **(A)** AE-Free time in Stable CF subjects separated into IL-1β-, TNFα-, G-CSF-, IL-6-, and IL-8-High and -Low groups over a 60–day follow up period. Marker-high and -low groups were defined according to sputum concentration thresholds obtained from ROCs separating CF Stable and AE levels (*Supplemental Figure 3*). AE-Free time was defined as the number of days from sputum collection in Stable subjects until the date of their next AE. **(B)** AE-Free time in Stable CF subjects separated into IL-1β-, TNFα-, G-CSF-, IL-6-, and IL-8- High and -Low groups over a 365–day follow up period. Cox Proportional Hazards model used to calculate AE-free intervals and adjust for age, sex, BMI, FEV_1_, number of *F508del* mutations, presence of CF-related diabetes or pancreatic insufficiency, use of CFTR correctors/modulators, and microbiology for *P. aeruginosa, A. xylosoxidans, H. parainfluenzae, Methicillin-sensitive S. aureus, and Methicillin-resistant S. aureus. HR: Hazard ration; SE: Standard error*.

**Supplemental Figure 7.**
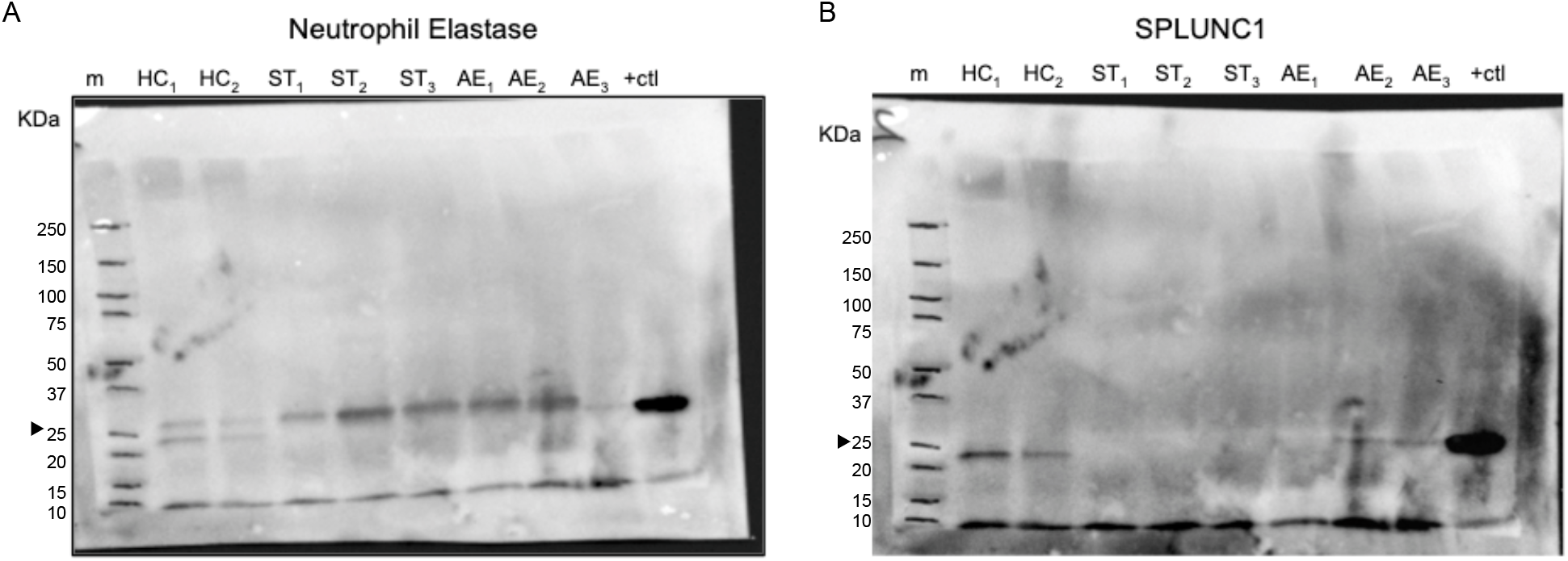
Full unedited Western blots for figure 5D. Membranes were initially probed for NE prior to stripping and re-probing for SPLUNC1. A) NE antibody: Mouse monoclonal anti-hELA2 raised against residues M1–N252 (1:3000, R&D systems).B) SPLUNC1 antibody: goat polyclonal hPLUNC1 antibody raised against residues Q20 – V256 of hPLUNC1 (1:3000, R&D systems), a secondary anti-goat HRP. HC: *Healthy Control; ST: Stable CF; AE: CF exacerbation; m: marker; +ctl: positive control*.

## Data Availability

data

## ACKNOWLEDGMENTS

This work was supported by The National Institutes of Health & National Heart, Lung, and Blood Institute (USA) through grants NIH R01-HL081160 and R21-AI083475 (LC), NIH T32-HL007778 and K01-HL125514-01 (CB), the Cystic Fibrosis Foundation through its Fifth Year Clinical Fellowship Award, and the American Thoracic Society Foundation Unrestricted Research Award (CB). CF Foundation (RT); UK CF Trust (RT). We thank our patients, the medical staff at the Yale Adult Cystic Fibrosis Program, and Dr. Jonathan Koff, Director of the Adult CF Program, for their support and contributions to this project. We also thank Dr. Mehmet Kesimer from the University of North Carolina at Chapel Hill for his thoughtful review and contributions in the development of this manuscript.

## AUTHOR CONTRIBUTIONS

CB, NN, SK, LS, and CDC planned the project, designed and performed experiments, analyzed the data, and wrote the manuscript. TL and MN, contributed to the design and analysis of clinical data, and provided access to the UMN cohort samples. MW, and RT performed, and analyzed experiments related to NE densitometry and activity in sputum. GC generated the sputum collection protocol for the Yale cohort and facilitated access to banked human samples. LC, MS, JLG, ME, and GC contributed to the design and analysis of all experiments, and the final manuscript. MDS provided biostatistics support and analyzed all experiments. All authors reviewed, revised, and approved the manuscript for submission.

This work was supported by NIH R01-HL081160 and R21-AI083475 (LC), NIH T32-HL007778, NIH/NHLBI K01-HL125514-01, and Cystic Fibrosis Foundation’s Fifth Year Clinical Fellowship (CB), American Thoracic Society Foundation Unrestricted Research Award (CB), CF Foundation (RT); UK CF Trust (RT).

